# Six-month pulmonary impairment after severe COVID-19: a prospective, multicenter follow-up study

**DOI:** 10.1101/2021.03.29.21254151

**Authors:** Paola Faverio, Fabrizio Luppi, Paola Rebora, Sara Busnelli, Anna Stainer, Martina Catalano, Luca Parachini, Anna Monzani, Stefania Galimberti, Francesco Bini, Bruno Dino Bodini, Monia Betti, Federica De Giacomi, Paolo Scarpazza, Elisa Oggionni, Alessandro Scartabellati, Luca Bilucaglia, Paolo Ceruti, Denise Modina, Sergio Harari, Antonella Caminati, Maria Grazia Valsecchi, Giacomo Bellani, Giuseppe Foti, Alberto Pesci

## Abstract

**Background and objective:** Long-term pulmonary sequelae following SARS-CoV-2 pneumonia are not yet confirmed, however preliminary observations suggests a possible relevant clinical, functional and radiological impairment. The aim of this study was to identify and characterise pulmonary sequelae caused by SARS-CoV-2 pneumonia at 6-month follow-up.

**Methods:** In this multicenter, prospective, observational cohort study, patients hospitalised for SARS-CoV-2 pneumonia and without prior diagnosis of structural lung diseases were stratified by maximum ventilatory support (“oxygen only”, “continuous positive airway pressure (CPAP)” and “invasive mechanical ventilation (IMV)”) and followed up at 6 months from discharge. Pulmonary function tests and diffusion capacity for carbon monoxide (DLCO), 6 minutes walking test, chest X-ray, physical exam and modified Medical Research Council (mMRC) dyspnoea score were collected.

**Results:** Between March and June 2020, 312 patients were enrolled (83, 27% women; median [IQR] age 61.1 [53.4,69.3] years). The parameters that showed the highest rate of impairment were DLCO and chest-X-ray, in 46% and 25% of patients, respectively. However, only a minority of patients reported dyspnoea (31%), defined as mMRC ≥ 1, or showed a restrictive ventilatory defects (9%). In the logistic regression model, having asthma as comorbidity was associated with DLCO impairment at follow-up, while prophylactic heparin administration during hospitalisation appeared as a protective factor. Need for invasive ventilatory support during hospitalisation was associated with chest imaging abnormalities.

**Conclusion:** DLCO and radiological assessment appear to be the most sensitive tools to monitor patients with COVID-19 during follow-up. Future studies with longer follow-up are warranted to better understand pulmonary sequelae.

**Summary at a glance:** DLCO and radiological assessment are the most sensitive tools to monitor COVID-19 patients at 6-month follow-up. Invasive ventilatory support is a risk factor for detection of radiological abnormalities, but not for DLCO impairment, at follow-up. Whileuse of prophylactic heparin acts as a protective factor on the development of DLCOimpairment.

## Introduction

The Coronavirus disease 2019 (COVID-19) pandemic, caused by the severe acute respiratory syndrome coronavirus 2 (SARS-CoV-2) and initiated in Wuhan (China) in December 2019, has expanded dramatically throughout the world during the last year.^1^ Pneumonia and acute respiratory distress syndrome (ARDS) are frequent manifestations of COVID-19; its pathogenic mechanisms are not entirely known and patients develop various degrees of respiratory failure, ranging between oxygen therapy, non-invasive ventilation (NIV) and invasive mechanical ventilation (IMV).

Some studies identified ARDS-like lesions, characterized by an inflammatory reaction in early-phase,^2^ leading to fibrotic sequelae or to the development of pulmonary emphysema, evolving to lung function impairment. Prior experience from H1N1 influenza pneumonia and severe acute respiratory syndrome (SARS) showed that long-term pulmonary fibrosis developed in up to 10% and 4.6% of patients with ARDS-like forms, respectively.^3,4^ Some more recent observations also highlighted the association between excessive distention of pulmonary parenchyma during IMV, leading to baro-volutrauma, and the development of post-ARDS pulmonary fibrosis.^5,6^

These previous observations may suggest a potentially relevant impact of pulmonary sequelae after severe COVID-19, however preliminary reports after 3 months of follow-up of these patients showed conflicting results,^7-9^ suggesting the importance to continue the follow-up of these patients. Furthermore, some of the short-term follow-up pulmonary sequelae, including ground glass opacities and atelectasis, require longer observation to assess whether they will be irreversible and its potential impact on pulmonary function. Furthermore, results from large multicenter prospective cohort studies with 6 and 12 months follow-up are in the majority of cases still ongoing.^10^

This study aims to identify and characterize pulmonary sequelae, in patients hospitalized for SARS-CoV-2 pneumonia, at 6 months follow-up after hospital discharge, and to evaluate their association with the maximum ventilatory support received during hospitalization.

## Materials and Methods

### Study design and participants

In this multicenter, prospective, observational cohort study, we enrolled consecutive patients hospitalized for laboratory-confirmed SARS-CoV-2 pneumonia between March and June 2020 in 7 hospitals in Lombardy, a region of Northern Italy populated by about 10 million people: San Gerardo Hospital, Monza; G. Salvini Hospital, Garbagnate Milanese; San Giuseppe Hospital, Milan; Spedali Civili, Brescia; Ospedale Civile, Vimercate; Ospedale Maggiore, Crema; Ospedale Maggiore, Cremona. Patients were followed up at 6 months from discharge to evaluate the presence of pulmonary sequelae with clinical evaluation, complete pulmonary function tests (PFTs) including plethysmography and diffusion capacity for carbon monoxide (DLCO) with single-breath technique, 6-minute walking test (6MWT) and chest X-ray. Clinical evaluation included the collection of a dyspnea score (Modified Medical Research Council (mMRC) scale) and lung auscultation to detect the presence of pathologic lung sounds.

Inclusion and exclusion criteria are summarized in Table 1.

**Table 1.** Inclusion and exclusion criteria.

| **INCLUSION CRITERIA** |
| --- |
| Age between 18 and 80 years |
| Diagnosis of SARS-CoV-2 infection by positive polymerase chain reaction (PCR) on nasal-pharyngeal swab or on bronchoalveolar lavage in case of double negative swab (GeneXpert® Cepheid; InGenius® Elitech; Abbott Real-Time SARS-CoV-2 assay Abbott; SARS-CoV-2 Plus ELITE MGB® Assay Elitech; Allplex™ SARS-CoV-2 Assay Arrow Diagnostics) |
| Clinical/instrumental signs of interstitial pneumonia and acute respiratory failure ($\text{PaO}_2/\text{FiO}_2 < 300$ in room air) on hospital admission |
| Written informed consent |
| Patients were discharged at home or in another hospital facility |
| **EXCLUSION CRITERIA** |
| Severe renal failure, defined as a glomerular filtration rate lower than 30ml/min |
| New York Heart Association (NYHA) class IV (unable to carry on any physical activity without discomfort) |
| Pregnancy or breastfeeding |
| Bacterial and/or fungal pulmonary superinfection during hospital stay |
| Prior diagnosis of chronic obstructive pulmonary disease, pulmonary emphysema, pulmonary fibrosis or bronchiectasis. Prior diagnosis of asthma and obstructive sleep apnea syndrome were not excluded since they do not cause permanent and irreversible radiological and/or functional impairment. |

Footnotes: $\text{PaO}_2/\text{FiO}_2$ = ratio of arterial oxygen partial pressure ( $\text{PaO}_2$ ) to fractional inspired oxygen ( $\text{FiO}_2$ )

Patients were stratified according to the maximum oxygen/ventilatory support received during hospital stay: 1) oxygen therapy alone; 2) continuous positive airway pressure (CPAP); 3) invasive mechanical ventilation (IMV). CPAP and IMV were applied according to the position papers on the management of respiratory failure in patients with COVID-19.^11^

This study received Ethics Committee approval (ASST Monza, 3389, May 21^st^ 2020) and was registered on clinicaltrial.gov (ClinicalTrials.gov Identifier: NCT04435327). The study is composed of two follow-up visits at 6 months and 1 year from hospital discharge. In this manuscript we report results from the 6-months visit. All patients provided written informed consent at the time of enrolment. The study is reported according to STROBE guidelines.^12^

### Procedures

PFTs and DLCO measurements were performed according to the American Thoracic Society (ATS)/European Respiratory Society (ERS) standardization using a dry spirometer.^13,14^ PFT parameters were expressed as absolute and percentage of a theoretical value calculated by Global Lung Function 2012 equations.^15^ 6MWT was performed according to the guidelines recommended by the ATS.^16^ The lower limits of normal for distance walked in healthy men and women were calculated according to the equation created by Enright and colleagues.^17^ Chest X-rays were evaluated by the pulmunologist and radiologist in charge for the presence of parenchymal abnormalities (reticulations, ground glass opacities and/or consolidation) and their extension (upper, mid, lower region for each lung). Lung auscultation was performed by a pulmunologist and the presence of pathological breath sounds (crackles, “velcro” crackles, wheezing, rhonchi, squawks and rales) were reported.

### Outcomes

The primary endpoint of the study was DLCO impairment (DLCO% <80% of predicted) evaluated at 6 months from hospital discharge.

The secondary endpoints of the study were also assessed at 6 months from hospital discharge and were: 1) Vital Capacity (VC), Forced Vital Capacity (FVC), Tiffeneau Index (FEV1/FVC ratio), Forced Expiratory Volume in the 1st second (FEV1), Total Lung Capacity (TLC) and Residual Volume (RV) alterations; 2) dyspnea evaluated through mMRC scale; 3) pathological lung sounds at chest auscultation; 4) radiological alterations on chest X-ray; and 5) variation from the expected of the normal distance walked on 6MWT.

### Statistical analysis

A sample size of 360 patients was calculated to provide 80% power to detect an increase from 2% (patients who received oxygen therapy alone) to 10% (patients who received CPAP or IMV) in the percentage of pulmonary sequelae at 1 year after hospital discharge with a logistic regression model and a statistical significance of 0.05. Assuming a 10% rate of drop-out, the sample size was increased to 400 patients, equally distributed in each of the three arms.

In the descriptive analysis qualitative variables have been summarized by counts and percentages, while quantitative characteristics by quartiles. Patients characteristics of the groups identified by maximum ventilatory support received were compared by Fisher exact test and Kruskal-Wallis rank sum test, as appropriate. In order to evaluate the association between maximum ventilatory support and pulmonary sequelae a logistic regression model was applied adjusting for predefined variables: age, gender, body mass index (BMI), cardiovascular diseases, diabetes, asthma, and treatment during hospital stay with systemic steroids or prophylactic heparin. Interactions were investigated and included in the model if statistically significant (p-value<0.05). A sensitivity analysis included also smoke as potential confounder. The model was fitted on the primary end-point (DLCO impairment) and on radiological alterations. Results were reported as odds ratio (OR) with 95% confidence interval (CI). The analyses were performed in R (version 4.0.4).

## Results

### Study population

In the study period, 420 consecutive hospitalized patients with SARS-CoV-2 pneumonia were screened for study participation. Out of the 420, 312 (83, 27% women; median [IQR] age 61.1 [53.4,69.3] years) met inclusion and exclusion criteria, provided consent and, thus, were enrolled in the final cohort and were stratified as follows: 71 patients in the “oxygen alone” group, 144 patients in the “CPAP” group and 97 patients in the “IMV” group, Figure 1. The baseline clinical features of the study population stratified by maximum oxygen/ventilatory support are shown in Table 2. The majority of patients were never smokers, with no differences between the study groups. The most frequently encountered comorbidities were obesity (34%), hypertension (29%), cardiovascular diseases (22%) and diabetes (14%). The majority of patients showed only one or the absence of comorbidities (78%). In regards to treatments received during hospitalization for COVID-19, patients in the “oxygen alone” group received significantly less specific treatments compared to the other groups. Pulmonary thromboembolism and deep vein thrombosis, two possible complications of COVID-19, were reported in 4.8% and 1.3% of patients, respectively, with no differences between groups.

**Table 2.** Demographics and clinical characteristics of study cohort.

| **DEMOGRAPHICS** |  |  |  |  |
| --- | --- | --- | --- | --- |
|  | **OXYGEN ONLY**<br>**(N=71)** | **CPAP**<br>**(N=144)** | **IMV**<br>**(N=97)** | **p** |
| Age (years), median [IQR] | 61.1 [53.3, 71.9] | 61.1 [53.1, 67.6] | 60.8 [55.2, 68.2] | 0.600 |
| Female gender, N (%) | 31 (44) | 33 (23) | 19 (20) | 0.001 |
| BMI (kg/m<sup>2</sup>), median [IQR] | 27.5 [24.6, 31.4] | 28.7 [26.6, 31.3] | 28.1 [25.7, 31.0] | 0.234 |
| Smoking History, N (%) |  |  |  | 0.100 |
| No | 47 (84) | 83 (65) | 51 (65) |  |
| Active | 1 (2) | 8 (6) | 6 (8) |  |
| Prior | 8 (14) | 36 (28) | 21 (27) |  |
| **COMORBIDITIES** |  |  |  |  |
|  | **OXYGEN ONLY** | **CPAP** | **IMV** | **p** |
| Cardiovascular Diseases, N (%) | 12 (17) | 31 (22) | 25 (26) | 0.397 |
| Hypertension, N (%) | 22 (31) | 43 (30) | 26 (27) | 0.816 |
| Cerebrovascular Diseases, N (%) | 1 (1) | 3 (2) | 1 (1) | 0.857 |
| Asthma, N (%) | 9 (13) | 4 (3) | 4 (4) | 0.015 |
| OSAS, N (%) | 2 (3) | 3 (2) | 1 (1) | 0.674 |
| Chronic Kidney Diseases, N (%) | 4 (6) | 2 (1) | 3 (3) | 0.162 |
| Liver diseases, N (%) | 2 (3) | 3 (2) | 0 (0) | 0.200 |
| Diabetes, N (%) | 12 (17) | 20 (14) | 13 (13) | 0.799 |
| Autoimmune Diseases, N (%) | 1 (1) | 2 (1) | 0 (0) | 0.603 |
| Prior Cancer, N (%) | 6 (8) | 2 (1) | 6 (6) | 0.026 |
| N. of Comorbidities, N (%) |  |  |  | - |
| 0 | 24 (34) | 68 (47) | 39 (40) |  |
| 1 | 29 (41) | 44 (31) | 38 (39) |  |
| 2 | 11 (15) | 25 (17) | 16 (16) |  |
| ≥ 3 | 7 (10) | 7 (5) | 4 (4) |  |
| **TREATMENTS ASSOCIATED WITH COVID-19** |  |  |  |  |
|  | **OXYGEN ONLY** | **CPAP** | **IMV** | **p** |
| Systemic Steroid, N (%) | 18 (31) | 65 (56) | 43 (58) | 0.003 |
| Heparin, N (%) | 18 (31) | 55 (47) | 44 (59) | 0.005 |
| Tocilizumab, N (%) | 5 (9) | 17 (15) | 19 (25) | 0.031 |
| Remdesivir, N (%) | 1 (2) | 2 (2) | 12 (16) | <0.001 |
| Mucolytics, N (%) | 13 (22) | 35 (30) | 30 (41) | 0.084 |
| Hyperimmune Plasma, N (%) | 0 (0) | 1 (1) | 1 (1) | 1.000 |
| Lopinavir/Ritonavir, N (%) | 21 (36) | 76 (66) | 34 (46) | <0.001 |
| Hydroxychloroquine, N (%) | 42 (72) | 100 (88) | 59 (80) | 0.042 |

Footnotes: BMI= body mass index; CPAP= continuous positive airway pressure; IMV= invasive mechanical ventilation; IQR= interquartile range; OSAS= obstructive sleep apnea syndrome

**Figure 1.**
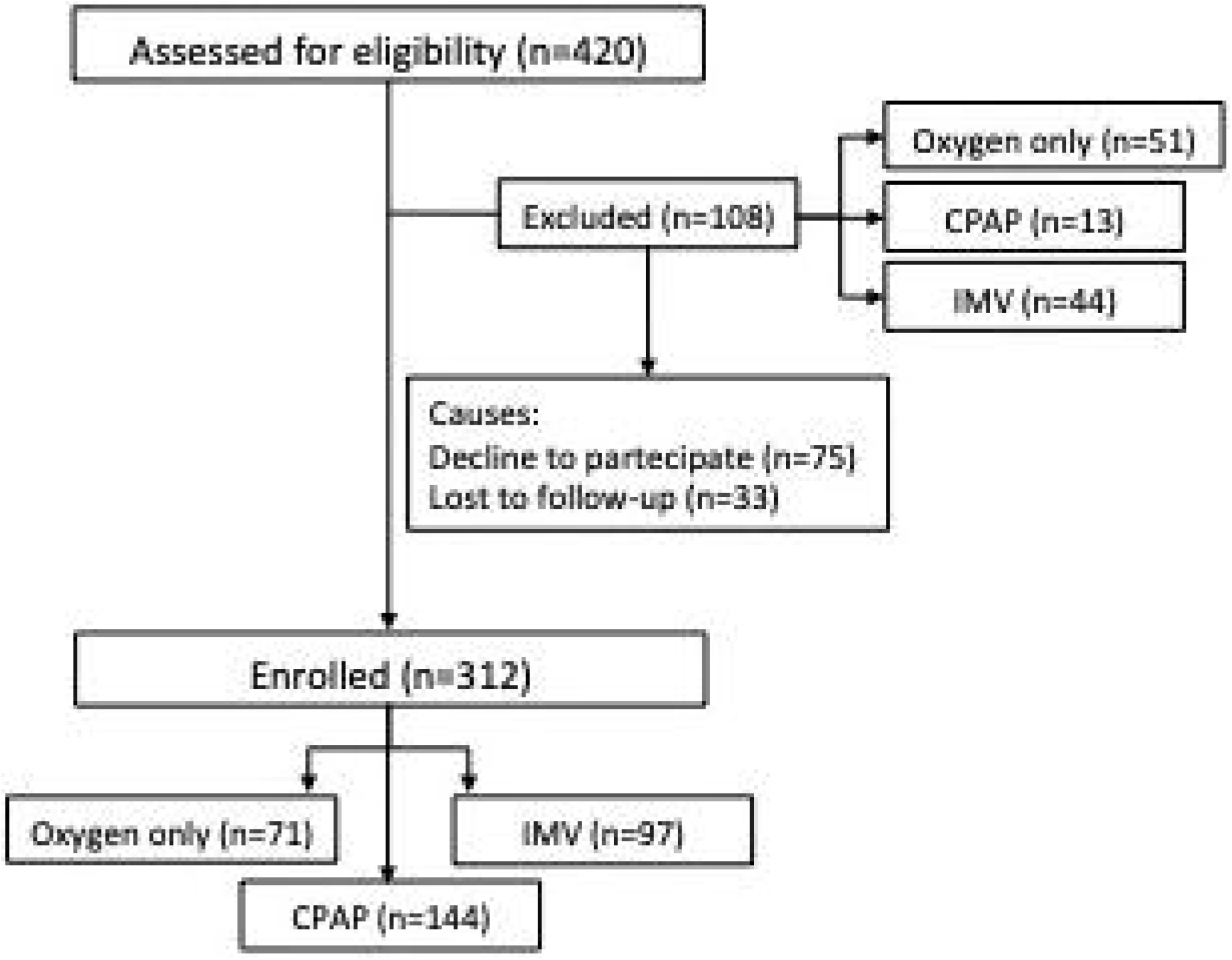
Study Flow Chart. Footnotes: CPAP= continuous positive airway pressure; IMV= invasive mechanical ventilation

### Evaluation of pulmonary sequelae

In regards to the presence of DLCO impairment (primary endpoint), we observed a statistically significant difference between groups with the highest prevalence of DLCO alteration in the “oxygen alone” (n= 40, 58%) and “IMV” group (n= 52, 54%) and the lowest in the “CPAP” group (n= 50, 36%), Table 3. However, patients in the “IMV” and “CPAP” group showed slightly more frequently moderate and severe DLCO impairment compared to the “oxygen alone” group, Figure S1 (Supplementary Information).

**Table 3.**
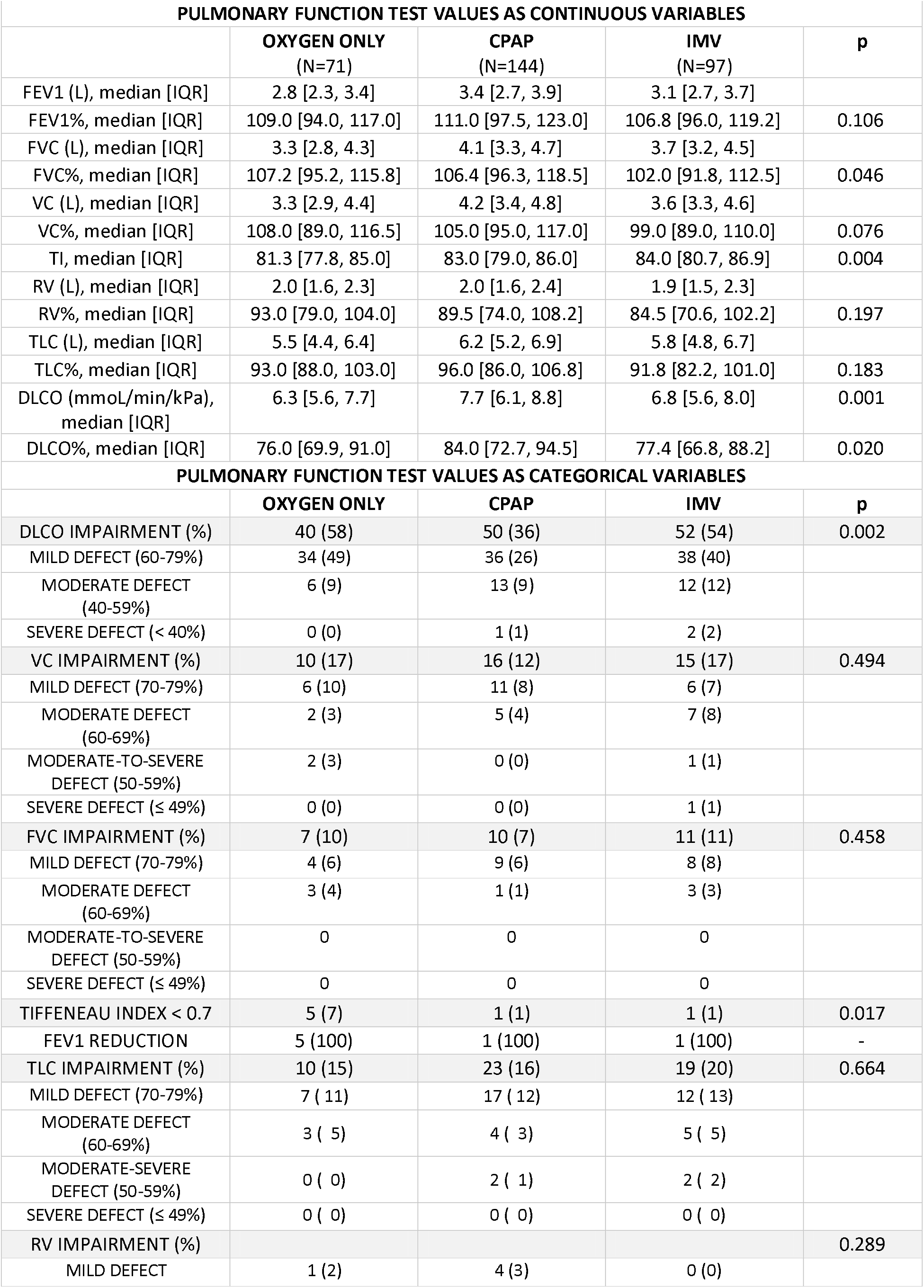
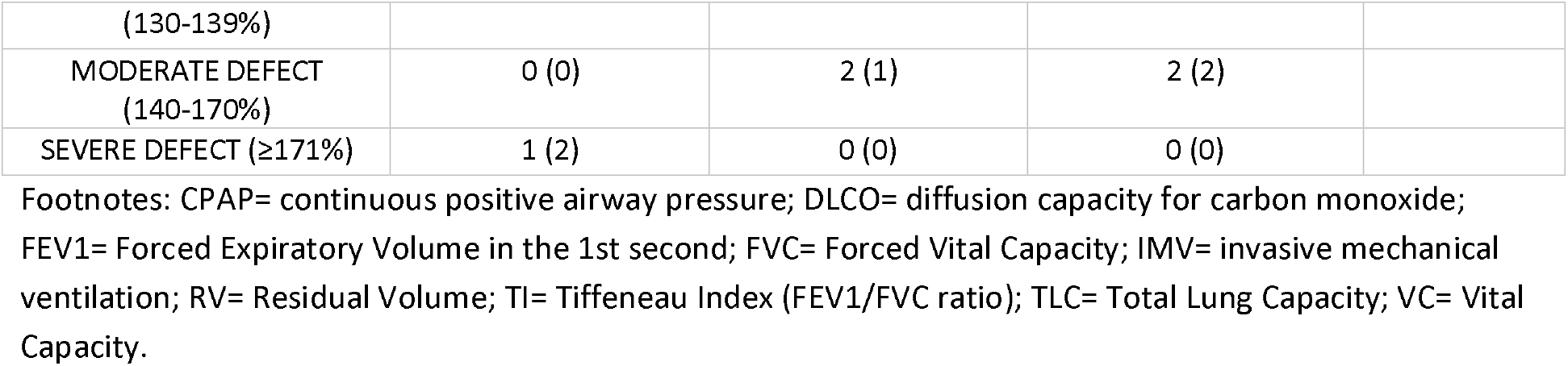
Pulmonary function tests at 6 months from hospital discharge.

No differences between groups were observed in the other parameters of PFTs, with the exception of FVC and Tiffeneau Index, Table 3. When considering FVC as a continuous variable, patients in the “IMV” group showed lower values compared to “CPAP” and “oxygen only” group (median [IQR] FVC% 102% [91.8-112.5], 106% [96.3-118.5] and 107% [95.2-115.8], respectively, p-value 0.046). However, when considering pathological values of FVC% (below 80% of predicted), only a minority of patients presented this condition (10, 7 and 11% of cases in the “oxygen only”, “CPAP” and “IMV” group, respectively) with no differences between groups. Therefore, only a minority of patients (28, 9%) showed a restrictive pattern. An obstructive pattern (defined as Tiffeneau Index < 0.7 with a concomitant reduction of FEV1 < 80%) was observed only in 7 (2.2%) patients, in the majority of cases in the “oxygen only” group (5 cases). Among the 7 cases with obstructive pattern one was active and two prior smokers and one had asthma as comorbidity.

Median distance walked at 6MWT ranged between 150 and 700 meters, with no differences between groups, Table 4. However, up to 46 (17%) of patients showed a distance walked lower than expected, again without differences between groups. No patients showed oxygen desaturation or required oxygen supplementation during the test.

**Table 4.**
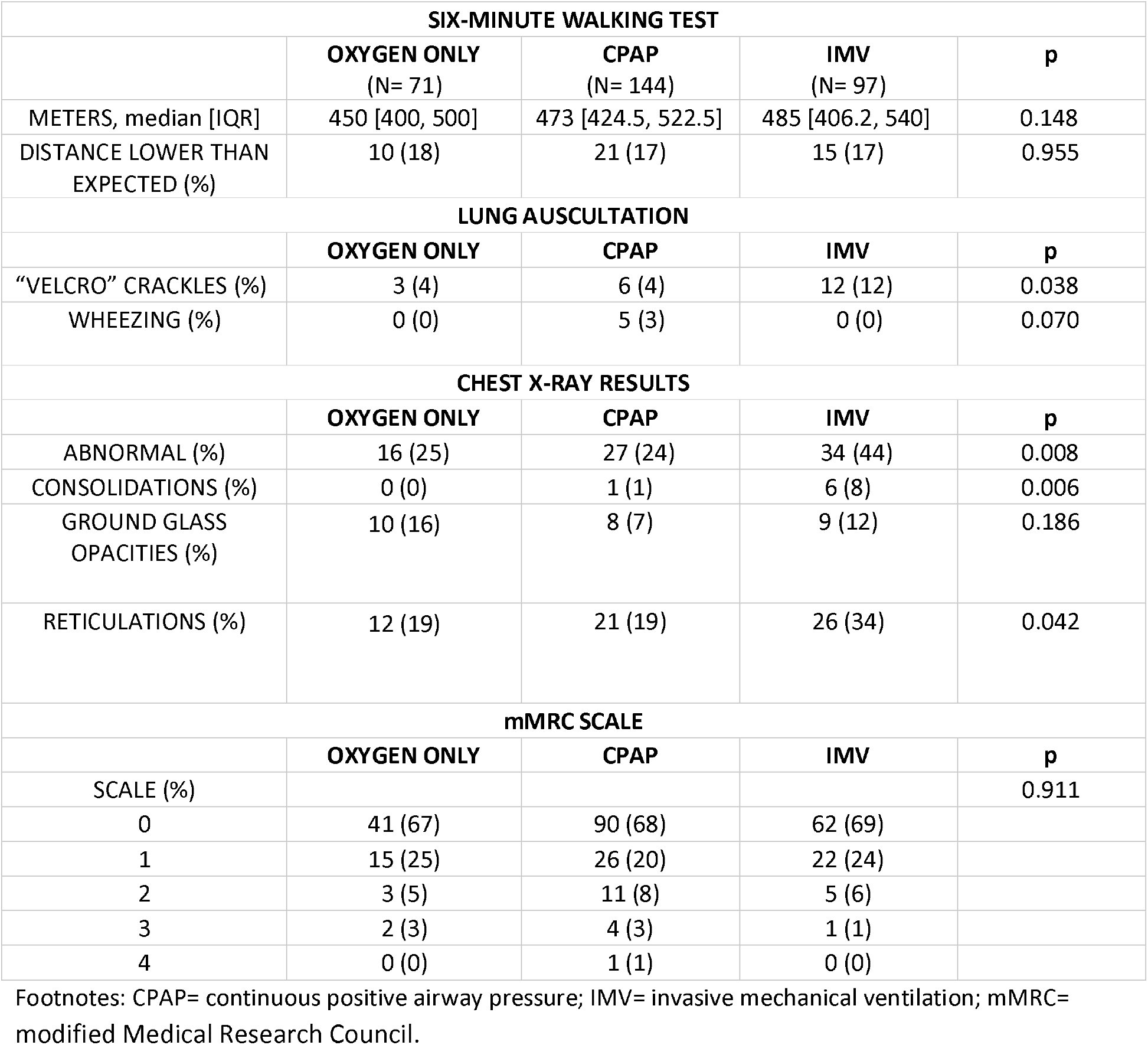
Six-minute walking test, lung auscultation, chest X-ray and mMRC scale results at 6 months from hospital discharge.

Characterizing the degree of dyspnoea reported by patients through the mMRC scale, the majority of patients (62%) were “not troubled by breathlessness except on strenuous exercise”, with no differences between groups, Table 4. Also at lung auscultation only a minority of patients, ranging from 4 to 12% according to study group, presented pathological sounds, mostly “velcro” crackles (21 cases) followed by wheezing (5 cases). “Velcro” crackles were significantly more frequent in the “IMV” group. Among the 21 cases with “velcro” crackles at physical exam, 10 (48%) showed abnormal chest X-ray (reticulations in 9 out of 10 cases), and 7 (33%) had DLCO impairment, of mild entity in all patients, but none showed restrictive pattern at PFTs. Three of the 5 cases who showed wheezing also had asthma, but none of them had an obstructive ventilatory defect.

Chest X-ray abnormalities were more frequently encountered in the “IMV” group (34, 44% of cases) compared to “CPAP” group (27, 24% of cases) and “oxygen only” group (16, 25% of cases), p value=0.008. The type of features more frequently observed were reticular in 59 patients (23%), ground glass opacities in 27 (11%) and consolidation in 7 (3%) of patients, Table 4.

After adjusting for demographics, comorbidities and treatments during hospital stay (Table 5), the “IMV” group showed higher odds of DLCO impairment with respect to the “oxygen only” group although the difference was not significant (OR=1.73, 95%CI: 0.75; 3.99). No significant difference in DLCO impairment was also observed among “CPAP” and “oxygen alone” (OR=0.72, 95%CI: 0.34; 1.54). Interestingly in subjects treated with prophylactic heparin the odds of DLCO alteration were halved (OR=0.45, 95%CI: 0.25; 0.83). Patients with asthma presented higher odds of altered DLCO (OR=4.86, 95%CI: 1.09; 21.68). No difference among the three groups was also observed in a sensitivity analysis including smoke in the model (not included in main analysis due to the low number of active smokers (n=15) and 16% of missing).

**Table 5.**
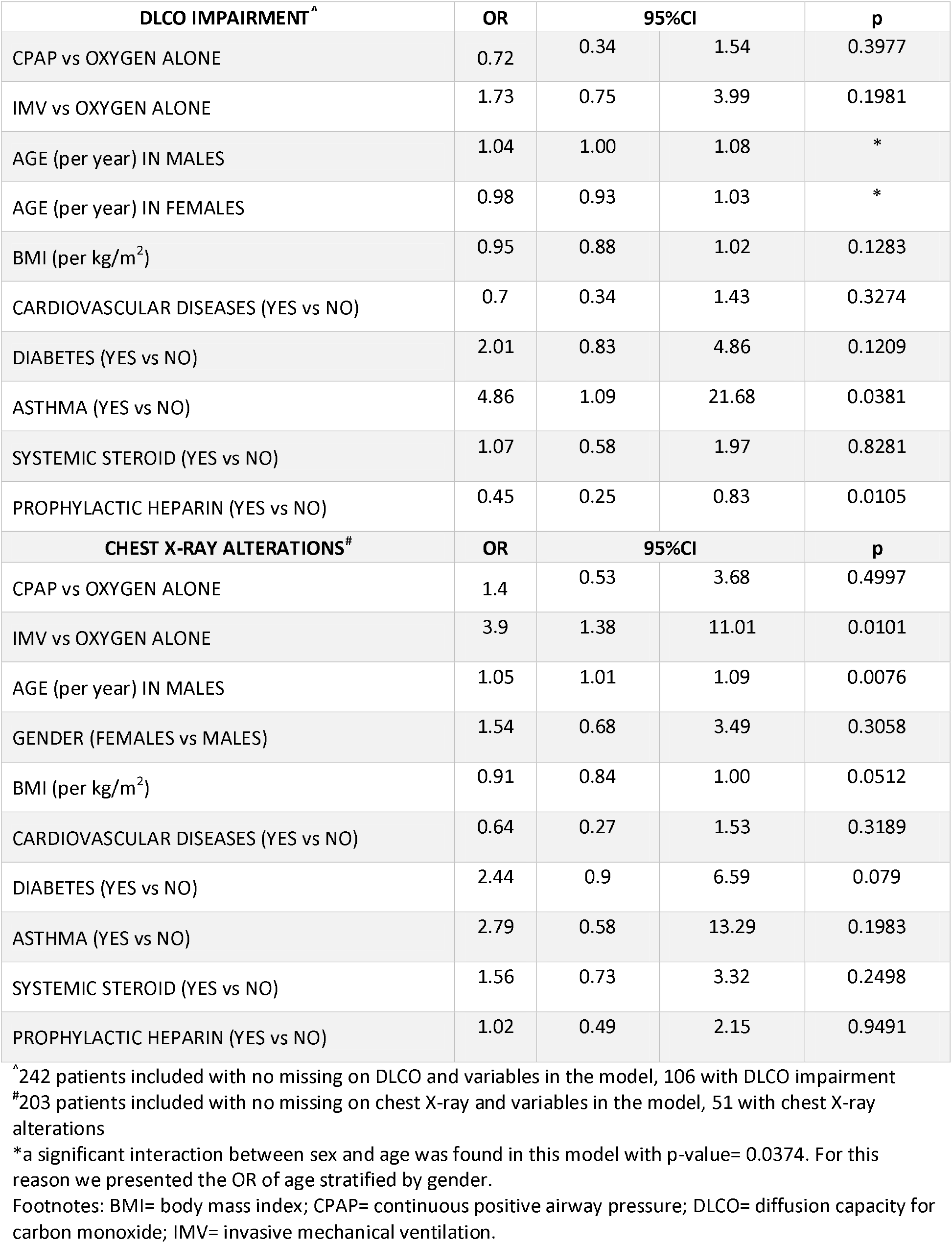
Multivariable logistic model results on the association between groups and DLCO or chest-X-ray impairment adjusted for prespecified variables.

The odds of radiological alterations on chest X-ray were higher in older patients (OR=1.05, 95%CI: 1.01;1.09) and in patients treated with “IMV” with respect to patients on oxygen alone (OR=3.9, 95%CI: 1.38; 11.01), while patients treated with CPAP did not showed significantly higher odds of radiological alterations.

## Discussion

We report the prevalence and degree of 6-month pulmonary sequelae in a cohort of 312 patients hospitalized for COVID-19 in 7 hospitals in Lombardy, the Italian region most populated and most severely affected by the pandemic so far.

We observed a considerable percentage of pulmonary sequelae when considering DLCO and chest X-ray. Up to 58% and 44% of patients according to study groups showed alterations of DLCO and chest X-ray, respectively. This was particularly evident in patients requiring IMV, but in the majority of cases the degree of impairment was mild.

Patients in the “oxygen only” group showed, together with the “IMV” group, the highest degree of DLCO impairment. We speculate that this may be partially related to the fact that the “oxygen only” group received significantly less specific treatments during hospitalization compared to the other groups, particularly in regards to systemic steroid (31% in the “oxygen only” group compared to 56% and 58% in the “CPAP” and “IMV” group, respectively) and prophylactic heparin (31% in the “oxygen only” group compared to 47% and 59% in the “CPAP” and “IMV” group, respectively). This could have influenced a slower recovery of normal lung anatomy and function at medium term follow-up. Thus, these preliminary observations need to be confirmed with longer follow-ups. After adjusting for potential confounders, such as treatment, DLCO impairment resulted higher for “IMV” group with respect to “oxygen only”, although with a not significant difference.

The lesson learned from interstitial lung diseases shows us that the DLCO is a more sensitive and earlier parameter to detect pulmonary dysfunction compared to 6MWT and ventilatory defects.^18^ Furthermore, DLCO can be altered both by parenchymal and pulmonary vascular diseases and COVID-19 may have a course characterized by an overlap between interstitial pneumonia with oedema and altered pulmonary perfusion with microthrombosis and macrothrombosis.^19^ Therefore, DLCO appears to be the most sensitive parameter among those available to monitor patients with COVID-19 during follow-up. Furthermore, on multivariable analysis the use of prophylactic heparin during hospitalization appeared to act as a protective factor on the development of DLCO impairment at 6 months follow-up. The potential beneficial effect of heparin during COVID-19 acute phase has been widely discussed, although randomized clinical trials are needed.^20^ The mechanisms involved include the antiviral and anti-inflammatory effect and the anticoagulant action on the hypercoagulability state associated with the disease. Our observations seem to suggest a prominent role of the vascular involvement during the acute phase of COVID-19 on the onset of DLCO impairment.

In our cohort need for invasive ventilatory support, that may be considered a proxy of disease severity, was a risk factor for the detection of chest imaging abnormalities at 6 months. The main alterations observed were reticulations and ground glass opacities. In particular, reticulations were significantly more frequent in patients who underwent IMV.

Our results nicely fit with a report on 736 patients from Wuhan followed up at 6 months after acute SARS-CoV-2 infection.^10^ This study showed a DLCO and radiological impairment in up to 56% and 54%, respectively, of patients requiring high-flow nasal cannula, non-invasive ventilation or IMV, and the severity of the acute disease was the major risk factor for the development of pulmonary sequelae.

Among the main strengths of our study we acknowledge: 1) the multicentric design, which included both university and non-university hospitals, allowed us to increase the study cohort and to enhance the generalizability of the results; 2) the selection criteria chosen excluded patients with pre-existing structural lung diseases and those who developed bacterial and/or fungal pulmonary infections during hospitalization, which may have caused PFT and/or radiological alterations not attributable to COVID-19.

Our study presents also some limitations: 1) the study visits were conducted during the second pandemic wave and this may have contributed to the lost to follow-up of some patients that were afraid of going to the hospital for medical visits, however the distribution of age and gender was similar among all patients recruited and patients actually visited; 2) data on the severity of radiological involvement during hospitalization, that may have had an impact on the development of pulmonary sequelae, were not collected, although the maximum ventilatory support needed by the patients gave us an hint about the severity of pneumonia.

In conclusion, up to 58% of patients with COVID-19, according to study group, present pulmonary sequelae, although of mild entity in the majority of cases, at 6-month follow-up. DLCO and radiological assessment appear to be the most sensitive tools to monitor patients with COVID-19 during follow-up. Need for invasive ventilatory support during hospitalization is a risk factor for detection of radiological abnormalities, but not for DLCO impairment, at follow-up. While use of prophylactic heparin acts as a protective factor on the development of DLCO impairment.

Future studies should evaluate the pulmonary sequelae developed by patients in the second and subsequent pandemic waves to assess the impact of standard-of-care therapies, such as steroid and heparin, that were not routinely used during the first wave. Furthermore, we await data on long-term sequelae with data at 1-year follow-up.

## Data Availability

Individual participant data referring to this article (i.e. text, tables and figures) will be made available upon reasonable request. The study protocol will be made available for researchers who provide a methodologically sound proposal. Proposals should be directed to

## Acknowledgments

We acknowledge Davide Gaudesi, PhD and Silvia Mori, PhD from Bicocca Clinical Research Organization (BiCRO) for their support in designing eCRF and project management.

We acknowledge that this research was partially supported by the Italian Ministry of University and Research (MIUR) - Department of Excellence project PREMIA (PREcision MedIcine Approach: bringing biomarker research to clinic).

We acknowledge Valentina Bonfanti, Pietro Curci, Giovanni Franco, Tommaso Passerella from University of Milano Bicocca for their support in follow-up organization and conduction.

---

**Figure S1. Degree of diffusion capacity for carbon monoxide (DLCO) impairment by maximum ventilatory support**.

## Bibliography

1 Guan WJ, Ni ZY, Hu Y, Liang WH, Ou CQ, He JX, Liu L, Shan H, Lei CL, Hui DSC, D. B, Li LJ, Zeng G, Yuen KY, Chen RC, Tang CL, Wang T, Chen PY, Xiang J, Li SY, Wang JL, Liang ZJ, Peng YX, Wei L, Liu Y, Hu YH, Peng P, Wang JM, Liu JY, Chen Z, Li G, Zheng ZJ, Qiu SQ, Luo J, Ye CJ, Zhu SY, Zhong NS. Clinical Characteristics of Coronavirus Disease 2019 in China. N. Engl. J. Med. 2020; 382: 1708–1720.

2 Tian S, Hu W, Niu L, Liu H, Xu H, Xiao S. Pulmonary Pathology of Early-Phase 2019 Novel Coronavirus (COVID-19) Pneumonia in Two Patients With Lung Cancer. J Thorac Oncol. 2020; 15(5):700–704.

3 Mineo G, Ciccarese F, Modolon C, Landini MP, Valentino M, Zompatori M. Post-ARDS pulmonary fibrosis in patients with H1N1 pneumonia: role of followup CT. Radiol Med. 2012; 117(2):185–200.

4 Zhang P, Li J, Liu H, Han N, Ju J, Kou Y, Chen L, Jiang M, Pan F, Zheng Y, Gao Z, Jiang B. Long-term bone and lung consequences associated with hospital-acquired severe acute respiratory syndrome: a 15-year follow-up from a prospective cohort study. Bone Res. 2020; 8:8.

5 Zhang R, Ying Pan Y., Fanelli V., Wu S, Luo AA, Islam D, Han B, Mao P, Ghazarian M, Zeng W, Spieth PM, Wang D, Khang J, Mo H, Liu X, Uhlig S, Liu M, Laffey J, Slutsky AS, Li Y, Zhang H. Mechanical Stress and the Induction of Lung Fibrosis via the Midkine Signaling Pathway. Am J Respir Crit Care Med. 2015; 192(3):315–23.

6 Cabrera-Benitez NE, Laffey JG, Parotto M, Spieth PM, Villar J, Zhang H, Slutsky AS. Mechanical Ventilation–associated Lung Fibrosis in Acute Respiratory Distress Syndrome: A Significant Contributor to Poor Outcome. Anesthesiology. 2014; 121(1): 189–198.

7 Zhao Y, Shangc Y, Song W, Li Q, Xie H, Xu Q, Jia J, Li L, Mao H, Zhou X, Luo H, Gao Y, Xu A. Follow-up study of the pulmonary function and related physiological characteristics of COVID-19 survivors three months after recovery. EClinicalMedicine. 2020;25:100463.

8 van Gassel R, Bels J, Raafs A, van Bussel BCT, van de Poll MCG, Simons SO, van der Meer LWL, Gietema HA, Posthuma R, van Santen S. High Prevalence of Pulmonary Sequelae at 3 Months after Hospital Discharge in Mechanically Ventilated Survivors of COVID-19. Am J Respir Crit Care Med. 2021; 1: 203(3):371–374.

9 van den Borst B, Peters JB, Brink M, Schoon Y, Bleeker-Rovers CP, Schers H, van Hees HWH, van Helvoort H, van den Bjoogaard M, Reijers MH, Prokop M, Vercoulen J, van den Heuvel M. Comprehensive health assessment three months after recovery from acute COVID-19. Clin Infect Dis. 2020; 21

10 Huang C, Huang L, Wang Y, Li X, Ren L, Gu X, Kang L, Guo L, Liu M, Zhou X, Luo J, Huang Z, Tu S, Zhao Y, Chen L, Xu D, Li Y, Li C, Peng L, Li Y, Xie W, Cui D, Shang L, Fan G, Xu J, Wang G, Wang Y, Zhong J, Wang C, Wang J, Zhang D, Cao B. 6-month consequences of COVID-19 in patients discharged from hospital: a cohort study. Lancet 2021; 397(10270):220–232.

11 Pfeifer M, Ewig S, Voshaar T, Randerath WJ, Bauer T, Geiseler J, Dellweg D, Westhoff M, Windisch W, Schönhofer B, Kluge S, Lepper PM Position Paper for the State-of-the-Art Application of Respiratory Support in Patients with COVID-19. Respiration. 2020;99(6):521-542.

12 von Elm E, Altman DG, Egger M, Pocock SJ, Gøtzsche PC, Vandenbroucke JP; STROBE Initiative.The Strengthening the Reporting of Observational Studies in Epidemiology (STROBE) Statement: guidelines for reporting observational studies. PLoS Med. 2007; 12(12):1495–9.

13 Graham BL, Steenbruggen I, Miller MR, Barjaktarevic IZ, Cooper BG, Hall GL, Hallstrand TS, Kaminsky DA, McCarthy K, McCormack MC, Oropez CE, Rosenfeld M, Stanojevic S, Swanney MP, Thompson BR. Standardization of Spirometry 2019 Update. An Official American Thoracic Society and European Respiratory Society Technical Statement. Am J Respir Crit Care Med. 2019; 200(8):e70–e88.

14 Graham BL, Brusasco V, Burgos F, Cooper BG, Jensen R, Kendrick A, MacIntyre NR, Thompson BR, Wanger J. 2017 ERS/ATS standards for single-breath carbon monoxide uptake in the lung. Eur Respir J. 2017 Jan; 49(1):1600016.

15 Quanjer PH, Stanojevic S, Cole TJ, Baur X, Hall GL, Culver BH, Enright PL, Hankinson JL, Ip MSM, Zheng J, Stocks J. Multi-ethnic reference values for spirometry for the 3-95-yr age range: the global lung function 2012 equations. Eur Respir J. 2012; 40: 1324–1343.

16 Holland AE, Spruit MA, Troosters T, Puhan MA, Pepin V, Saey D, McCormack MC, Carlin BW, Sciurba FC, Pitta F, Wanger J, MacIntyre N, Kaminsky DA, Culver BH, Revill SM, Hernandes NA, Andrianopoulos V, Camillo CA, Mitchell KE, Lee AL, Hill CJ, Singh SJ. Official European Respiratory Society/American Thoracic Society technical standard: field walking tests in chronic respiratory disease. Eur Respir J. 2014;44(6):1428–46.

17 Enright P L, Sherrill D L. Reference equations for the six-minute walk in healthy adults. Am J Respir Crit Care Med. 1998; 58(5 Pt 1):1384–7.

18 Ciancio N, Pavone M, Torrisi SE, Vancheri A, Sambataro D, Palmucci S, Vancheri C, Di Marco F. Contribution of pulmonary function tests (PFTs) to the diagnosis and follow up of connective tissue diseases. Multidiscip Respir Med. 2019; 14: 17.

19 Camporota L, Vasques F, Sanderson B, Barrett NA, Gattinoni L. Identification of pathophysiological patterns for triage and respiratory support in COVID-19. Lancet Res Med. 2020; 8(8):752–754.

20 Hippensteel JA, LaRiviere WB, Colbert JF, Langouët-Astrié CJ, Schmidt EP. Heparin as a therapy for COVID-19: current evidence and future possibilities. Am J Physiol Lung Cell Mol Physiol. 2020; 319(2):L211–L217.

