## Supplementary figures and images for "Six-month pulmonary impairment after severe COVID-19: a prospective, multicenter follow-up study"

### Figure S1

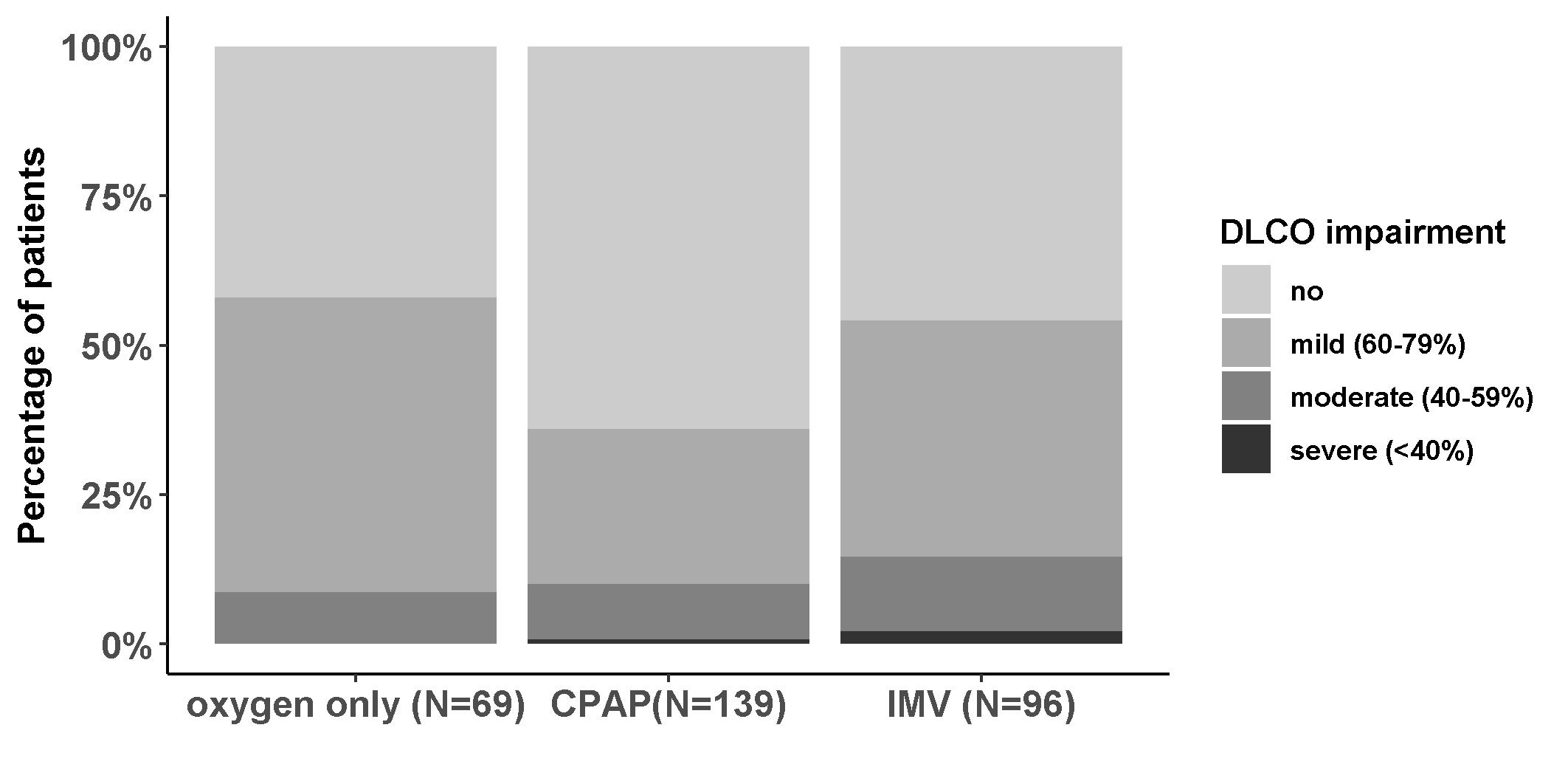
